# Enterprise healthcare physician services in Canada: an environmental scan

**DOI:** 10.1101/2022.04.22.22274184

**Authors:** Sheryl Spithoff, Lana Mogic

## Abstract

**Background:** Employers in Canada are increasingly offering physician services to their employees, often through third party workplace “enterprise healthcare” platforms. To date however, little work has been done to understand this method of organizing and delivering care.

**Objective:** To understand the nature, extent and implications of enterprise healthcare physician services in Canada.

**Methods:** We conducted structured internet and database searches to identify enterprise healthcare platforms that provided physician services and their public websites. To answer our research question, We extracted data from company websites and linked company documents as well as information from Mergent Intellect, a web-based application with business data on Canadian companies.

**Results:** We identified nine companies offering enterprise physician services to employees in Canada via 11 enterprise software platforms. According to company claims, over four million Canadian employees and their family members have access to enterprise physician services. All platforms offer virtual physician services and five also facilitate in person visits. Ten of the platforms provide primary care services and one offers only addiction medicine services. Four of the platforms offer to communicate and share information with an employee’s regular primary care provider. Five state they share aggregate or de-identified health data with employers.

**Conclusions:** Enterprise healthcare companies provide millions of Canadian employees and their families with rapid access to virtual physician services and, in some cases, in person care. These services may disrupt continuity of care (care by the same provider over time) and pose risks to employee privacy. As other Canadians do not have access to these services, enterprise healthcare is also introducing two-tiered healthcare across Canada potentially affecting the sustainability of the public healthcare system.

## Introduction

Employers in Canada are increasingly offering physician services to their employees, typically through a third party company (1–5). The third party companies enable virtual physician services through a workplace “enterprise healthcare” platform (Box 1). Some also facilitate in person physician care through the platforms (6). The companies claim the enterprise platforms improve access to care, leading to lower rates of employee absenteeism (7,8) and a good return on investment for employers (7–9). Some enterprise healthcare companies may share de-identified employee health data with employers (10–12). The companies state that data sharing allows employers to evaluate the health services they provide and identify ways to reduce employee absenteeism. Data sharing, however, also has implications for employee privacy and autonomy.

**Box 1.** Terms used in the article

Box 1. Terms used in the article
|  |  |
| --- | --- |
| Virtual care (Telemedicine) | Any interaction between patients and/or members of their circle of care (e.g. physicians, nurses), occurring remotely, using any forms of communication or information technologies with the aim of facilitating or maximizing the quality and effectiveness of patient care (20) |
| Enterprise healthcare | A method of delivering healthcare to employees, on behalf of the employer, using a third party’s proprietary software platform |
| Enterprise healthcare companies | The third-party companies contracted to provide enterprise healthcare services to employees, on behalf of employers |

Enterprise healthcare differs from the traditional model of healthcare in Canada where most people receive care from self-employed physicians working independently or in small groups in community clinics (13). These physicians either bill the provincial/territorial single-payer public health system – known as “Medicare” – or, much less commonly, bill patients directly (14–16). Until recently, these physicians, working in the traditional model of care, rarely provided virtual care; only three percent of Canadians reported a virtual care physician visit in 2018 (17). With the onset of the Covid-19 pandemic, and the introduction of public funding for virtual care, more physicians started offering these services (18). In the province of Ontario, for example, virtual care visits increased 56-fold to comprise over 70% of all visits between March 2020 and July 2020 (19).

Despite the apparent widespread access to enterprise healthcare in Canada, little work has been done to understand the extent and nature of this method of organizing and delivering physician services, nor the implications for employees and for the health system. Our research objective, therefore, was to gain an understanding of the nature, extent and implications of enterprise healthcare in Canada through a national environmental scan.

## Methods

We conducted structured internet searches between December 3^rd^, 2021 and April 1^st^, 2022 to identify enterprise platforms that provide physician services to company employees in Canada. The search terms used consisted of terms pertaining to the topic areas of virtual care, Canada, and enterprise. We ran each of the searches individually and examined the first 50 results looking for relevant findings. We compared our findings to a list we compiled when conducting a previous research study examining direct-to-patient virtual care in Canada to ensure our search terms were identifying all relevant platforms. We then identified the company that owned the platform.

### Inclusion/exclusion criteria

We included platforms if the company could be contracted by an employer to provide physician healthcare services through a proprietary software platform to employees living in Canada. We only included companies that provided physician services as these historically are part of Canadian Medicare, whereas many other clinical services (e.g., counseling) are not. We included companies with platforms that provided physician visits through both synchronous (e.g., phone calls, video chat, in person) and asynchronous communication (e.g., text communication). We excluded companies if they provided services solely to employees living outside of Canada or if they offered only non-physician services. We only included companies with platforms using the English language.

### Identification of documents

For each enterprise healthcare platform we included, we identified the platform’s public website and any linked websites or documents. We also searched Mergent Intellect, a publicly accessible, web-based application offering business data for a collection of US and Canadian private and public corporations (21). This database contains basic company information, as well as financial, industry, and executive details for over 1.6 million Canadian businesses. It offers information on corporate structures, including a list of key executives and a complete company family tree.

### Data extraction and analysis

We extracted information on the company’s corporate structure, company headquarters, countries of operation, ownership (publicly-traded or privately held), communication mode(s) (e.g., in person, text, phone, or video), primary care physician services, specialist services, payment for physician services (private or public), in person services, system navigator services, family coverage for services, sharing of data with employers and information on communication with an employee’s regular primary care provider. Where available, we also collected information on company claims about number of Canadian enterprise clients (employers) and number of enterprise members (employees and their family members) in Canada.

### Ethics

As we only used publicly available documents, we received a research ethics board exemption from Women’s College Hospital.

## Results

We identified 11 enterprise healthcare platforms offering physician services to employees in Canada. These enterprise care companies claim to provide services to more than 44,000 companies and 4.5 million employees and their family members (Table 1). Seven of the 11 companies are publicly-traded. Ten companies are based in Canada and one in the U.S. (Table 2). All companies offer virtual care physician services and five also arrange in person visits through their platform. Ten companies offer primary care and one company offers only addiction physician services. Seven provide coordinated access to specialist appointments and six provide a system navigator to help employees navigate the health system. Four platforms offer to provide information to an employee’s regular primary care provider and five state they provide employers with aggregate or de-identified employee health information.

**Table 1.** Companies providing enterprise physician services

| Platform name | Parent company | Claims about services provided |
| --- | --- | --- |
| ALAViDA | Lifespeak | - |
| Appletree | Appletree Medical Group | - |
| CloudMD | CloudMD Software & Services | 7200 companies (employers) |
| Dialogue | Dialogue Health Technologies | 25,000 plus companies (employers), 1.8 million members (employees and family members) |
| JungoHR | JungoHR | - |
| Kii | CloudMD Software & Services | - |
| Lifeworks | Telus | - |
| Maple | Maple Corporation | - |
| Meira Care | Meira Care | - |
| Teladoc | Teladoc Health | 12,000 plus companies (employers) |
| TELUS Health Virtual Care | Telus | 2.8 million members (employees and family members) |

**Table 2.** Description of companies providing enterprise physician services in Canada

|  |  | Number (%) |
| --- | --- | --- |
| Ownership | Privately-held | 4 (36%) |
|  | Publicly-traded | 7 (64%) |
| Headquarters | Canada | 10 (91%) |
|  | US | 1 (9%) |
| Funding of physician services | Public | -- |
|  | Private | -- |
|  | Not stated | 11 (100%) |
| Medical services | Primary care services | 10 (91%) |
|  | Addiction physician services only | 1 (10%) |
|  | Access to specialist care | 7 (64%) |
|  | System navigators | 6 (55%) |
|  | Family coverage | 5 (45%) |
| Modality | Virtual care | 11 (100%) |
|  | In person care | 5 (45%) |
| Access | 24/7 | 8 (73%) |
|  | Same day | 2 (22%) |
| Data sharing | With employee's regular primary care provider | 4 (36%) |
|  | With employers (aggregate or de-identified health data only) | 5 (45%) |

% = percentage

## Discussion

Our environmental scan provides insight into enterprise physician services in Canada. Enterprise healthcare companies provide millions of Canadian employees and their families with rapid access to virtual physician services and, in some cases, in person care. These services may disrupt continuity of care (care by the same provider over time) and pose risks to employee privacy. Further, as many people living in Canada do not have access to these services, the model is also introducing two-tiered healthcare broadly across Canada. This is likely to increase disparities in care and costs to the public health system.

Our analysis indicates that some employees may experience disruptions to continuity of care. Less than half of the platforms enable in-person physician visits, implying patients are expected to seek care from their regular care provider or the emergency department for complicated issues. As a result, employees are likely to experience a disruption to continuity of care (care by the same provider over time), a challenge for effective management of chronic and complex health conditions (6).

As some of these enterprise care companies state they share de-identified or aggregate patient data with employers and/or third parties, this method of delivering healthcare may present additional risks to employees. Although the risk of re-identification, is likely to be low if the data are de-identified or aggregated (1,22), sharing data with employers can enable other privacy-related harms such as surveillance and discrimination. For example, the data can be used to identify the characteristics of employees who are likely to have complex health conditions or who may have a high-risk pregnancy (10–12,23). Companies may use this information to improve services for these types of employees. They may also, however, use this information in job recruitment algorithms to ensure these individuals are not offered employment in the first place (24–26).

Enterprise healthcare is also introducing a two-tiered system, where millions of employees and family members have access to physician services not available to other Canadians. This may worsen health disparities. Employees of larger companies are often from a higher socio-economic stratum than other people living in Canada, and part of a group that already has better access to care (13), and better health outcomes (21–24). Further, studies show that a two-tiered system is likely to increase wait-times for those in the public system by luring physicians and other healthcare providers to the enterprise system (27–29).

Finally, enterprise healthcare may lead to “cream-skimming” (30,31). As mentioned above, many enterprise platforms do not offer in person assessments meaning that physicians working for these platforms can only treat simple medical problems. Patients who have complicated health issues are shunted to the emergency department or back to their regular primary care provider (28,32,33). This may increase the burden on the public health system (30,31). Recent research supports this concern. A Canadian study found that patients who had a virtual care visit with a commercial virtual care platform were more likely to have an emergency department visit that those who had a virtual care visit with their regular primary care provider (18).

### Limitations

Our study is limited by the fact that we relied on company documents to answer our research questions. Additionally, we were not able to find information on whether physician services were billed to the public system, or if companies paid for these services privately.

## Conclusion

Enterprise healthcare seeks to provide employees with rapid access to physician services. This method of organizing and delivering care, however, may also expose employees to harms, as well as pose threats to the sustainability of the public healthcare systems in Canada. More research is needed to further explore this method of delivering and funding physician services in Canada including how the system functions economically, who the system benefits and the implications for employees, other people in Canada and the publicly-funded health systems.

## Data Availability

All data produced in the present study are available upon reasonable request to the authors

## Acknowledgements

This project received funding from the Social Science and Humanities Council of Canada (SSHRC). Sheryl Spithoff receives funding from a New Investigator award from the Department of Family and Community Medicine, University of Toronto.

## References

1. CloudMD. CloudMD Announces Rapid Growth of Enterprise Health Solutions Division [Internet]. CloudMD Software & Services Inc. [cited 2021 Aug 30]. Available from: https://investors.cloudmd.ca/cloudmd-announces-rapid-growth-of-enterprise-health-solutions-division/

2. Shoppers Drug Mart. Shoppers Drug Mart and SilverCloud Health bring leading digital mental health solution to Canadian employers [Internet]. [cited 2021 Mar 30]. Available from: https://www.newswire.ca/news-releases/shoppers-drug-mart-and-silvercloud-health-bring-leading-digital-mental-health-solution-to-canadian-employers-840010994.html

3. Maple. Businesses [Internet]. [cited 2021 Mar 30]. Available from: https://www.getmaple.ca/business/

4. Teledact Inc. Gotodoctor.ca to provide on-demand virtual care services to frontline staff of Union Social Group restaurants [Internet]. [cited 2021 Aug 11]. Available from: https://www.newswire.ca/news-releases/gotodoctor-ca-to-provide-on-demand-virtual-care-services-to-frontline-staff-of-union-social-group-restaurants-806856625.html

5. PRLive. Digital Healthcare Creating a Potential $634.9 Billion Opportunity [Internet]. Baystreet.ca. 2021 [cited 2021 Mar 30]. Available from: /contact/

6. Workplace Health [Internet]. Appletree Medical Group; [cited 2022 Jul 19]. Available from: https://appletreemedicalgroup.com/wp-content/uploads/2021/05/Workplace-Health-WSIB-2021.pdf

7. Virtual Care | TELUS Health [Internet]. TELUS. [cited 2022 Apr 13]. Available from: https://www.telus.com/en/health/organizations/group-health-benefits/employers/virtual-care

8. The Maple experience [Internet]. Maple. [cited 2022 Apr 13]. Available from: https://www.getmaple.ca/business/experience/

9. Normandeau M. What makes Dialogue a valued investment in organizational wellness? [Internet]. [cited 2022 Apr 13]. Available from: https://www.dialogue.co/en/blog/dialogue-valued-investment-organizational-wellness

10. Companies Are Using Big Data To Track Employee Health And Pregnancies [Internet]. Popular Science. 2016 [cited 2022 Apr 10]. Available from: https://www.popsci.com/companies-use-big-data-to-track-employee-health-and-pregnancies/

11. Board TE. Opinion | Protecting Employees’ Health Data. The New York Times [Internet]. 2016 Mar 26 [cited 2022 Apr 10]; Available from: https://www.nytimes.com/2016/03/27/opinion/sunday/protecting-employees-health-data.html

12. Mark Witte: How to make employee health data work for your business and employees [Internet]. HRreview. 2019 [cited 2022 Apr 10]. Available from: https://www.hrreview.co.uk/analysis/mark-witte-how-to-make-employee-health-data-work-for-your-business-and-employees/116608

13. Martin D, Miller AP, Quesnel-Vallée A, Caron NR, Vissandjée B, Marchildon GP. Canada’s universal health-care system: achieving its potential. Lancet. 2018;391(10131):1718–35.

14. The illegality of private healthcare in Canada [CMAJ - March 20, 2001] [Internet]. [cited 2022 Apr 10]. Available from: https://www.nlc-bnc.ca/eppp-archive/100/201/300/cdn_medical_association/cmaj/vol-164/issue-6/0825.asp

15. Contandriopoulos D, Law MR. Policy changes and physicians opting out from Medicare in Quebec: an interrupted time-series analysis. CMAJ. 2021 Feb 16;193(7):E237–41.

16. Flood CM. Two-tier healthcare after Cambie - Colleen M. Flood, 2021. Healthcare Management Forum [Internet]. 2021 Mar 2 [cited 2022 Feb 1]; Available from: http://journals.sagepub.com/doi/full/10.1177/0840470421994304

17. Connecting patients for better health: 2018 [Internet]. Canada Health Infoway; 2018 [cited 2022 Apr 7]. Available from: https://www.infoway-inforoute.ca/en/component/edocman/3564-connecting-patients-for-better-health-2018/view-document?Itemid=0

18. Lapointe-Shaw L, Salahub C, Bhatia RS, Desveaux L, Glazier RH, Hedden L, et al. Characteristics and healthcare use of patients attending virtual walk-in clinics: a cross-sectional analysis [Internet]. medRxiv; 2022 [cited 2022 Mar 12]. p. 2022.02.28.22271640. Available from: https://www.medrxiv.org/content/10.1101/2022.02.28.22271640v1

19. Glazier RH, Green ME, Wu FC, Frymire E, Kopp A, Kiran T. Shifts in office and virtual primary care during the early COVID-19 pandemic in Ontario, Canada. CMAJ. 2021 Feb 8;193(6):E200–10.

20. Shaw J, Jamieson T, Agarwal P, Griffin B, Wong I, Bhatia RS. Virtual care policy recommendations for patient-centred primary care: findings of a consensus policy dialogue using a nominal group technique. J Telemed Telecare. 2018 Oct;24(9):608–15.

21. Mergent Intellect [Internet]. Toronto Public Library. [cited 2022 Jan 17]. Available from: https://www.torontopubliclibrary.ca/detail.jsp?Entt=RDMEDB0188&R=EDB0188

22. Big Data in Healthcare: How Employers, Providers, and Brokers Are Using Healthcare Analytics [Internet]. [cited 2021 Mar 30]. Available from: https://www.artemishealth.com/blog/big-data-in-healthcare-how-employers-providers-and-brokers-are-using-healthcare-analytics

23. Landi H. Modern Health rolls out data tool for employers to better pinpoint workers’ mental health needs [Internet]. Fierce Healthcare. 2021 [cited 2022 Apr 10]. Available from: https://www.fiercehealthcare.com/tech/modern-health-rolls-out-data-tool-for-employers-as-companies-ramp-up-efforts-to-address

24. Regan PM, Jesse J. Ethical challenges of edtech, big data and personalized learning: twenty-first century student sorting and tracking. Ethics Inf Technol. 2019 Sep 1;21(3):167–79.

25. O’Neil C. Weapons of Math Destruction: How Big Data Increases Inequality and Threatens Democracy. 1st edition. New York: Crown; 2016. 272 p.

26. Ebeling MFE. Healthcare and Big Data: Digital Specters and Phantom Objects. 1st ed. 2016 edition. Vol. p.43. New York: Palgrave Macmillan; 2016.

27. Duckett SJ. Private care and public waiting. Aust Health Rev. 2005 Feb;29(1):87–93

28. Lee SK, Rowe BH, Mahl SK. Increased Private Healthcare for Canada: Is That the Right Solution? Healthc Policy. 2021 Feb;16(3):30–42.

29. Cheng TC, Kalb G, Scott A. Public, private or both? Analyzing factors influencing the labour supply of medical specialists. Canadian Journal of Economics/Revue canadienne d’économique. 2018;51(2):660–92.

30. Cheng TC, Haisken-DeNew JP, Yong J. Cream skimming and hospital transfers in a mixed public-private system. Soc Sci Med. 2015 May;132:156–64.

31. Duckett S. Commentary: The Consequences of Private Involvement in Healthcare – The Australian Experience. Healthc Policy. 2020 May;15(4):21–5.

32. Davidson A. Parallel Payers, Privatization and Two-Tier Healthcare in Canada. HealthcarePapers [Internet]. 2008 May 15 [cited 2021 Aug 30];8(3). Available from: https://www.longwoods.com/content/19793/healthcarepapers/parallel-payers-privatization-and-two-tier-healthcare-in-canada

33. Davidson A. Under the Radar: Stealth Development of Two-Tier Healthcare in Canada. Healthc Policy. 2006 Jul;2(1):25–33.

